# High-risk HPV Prevalence and Distribution in Cervical Cancer Screening: Extended Genotyping in Correlation with Cytology and p16/Ki67 Dual Stain Results

**DOI:** 10.1101/2025.02.08.25321924

**Authors:** Martyna Trzeszcz, Karolina Mazurec, Maciej Mazurec, Robert Jach, Agnieszka Halon

**Author notes:** **Corresponding author:** Martyna Trzeszcz, MD, PhD, Division of Pathology and Clinical Cytology, University Hospital in Wroclaw, Borowska 213 St., 50-556 Wroclaw, Poland. **Conflicts of Interest** M.T. has given lectures sponsored by the Hologic company, received coverage for travel and accommodation costs. R.J. has given a lecture sponsored by the Roche company. No disclosures were reported by the other authors.

## Abstract

The prevalence of high-risk human papillomavirus (HR-HPV) in cytology combined with p16/Ki67 status using limited and two types of extended HPV genotyping has not yet been described. A total of 32,724 screening tests results between 2015-2024 were included. The overall HR-HPV-positivity rate was 15.0%. The HR-HPV prevalence in limited genotyping group was 13.9%, in extended genotyping 1 (17.8%), in extended genotyping 2 (17.2%), with statistically significant difference in the proportions of positive/negative cases (p<0.0001). No statistically significant difference was noted between extended genotyping groups (p=0.706). Extended genotyping 1: the highest p16/Ki67-positivity was observed for HR-HPV 33/58 (100.0%) and 31 (58.8%), the lowest for HR-HPV 45 (18.2%), 18 (25.0%) and 59/56/66 (28.9%). Extended genotyping 2: the highest p16/Ki67-positivity was for HR-HPV 16 (66.7%) and 31/33/52/58 (58.8%). A combined approach, implementing new technologies, could help healthcare providers in more informed decisions about patient care, supporting the prevention of cervical precancer and cancer.

**Statement of Significance:** The transition to HPV-based cervical cancer screening is progressively advancing. By investigating of HR-HPV prevalence and distribution in limited or extended genotyping, cytology and p16/Ki67 dual-stain, our study aimed to support evidence-based decision-making processes and help guide of medical interventions of public health policies targeted to incorporation of new technologies.

## Introduction

Human papillomavirus (HPV) infection is recognized as one of the most common sexually transmitted infections globally, with over 100 identified genotypes [1]. While most HPV infections are transient and asymptomatic, persistent infection with HR-HPV genotypes, with the predominance of HPV 16 and 18, is a well-known major risk factor for the development of cervical cancer, which is the fourth most prevalent cancer in women worldwide [2–4].

Previous studies have reported varying prevalence rates of HPV infection among women, highlighting the need for updated and comprehensive data from different regions [5,6]. The prevalence of HPV infection varies geographically and demographically, and is influenced by factors such as sexual behavior, socioeconomic status, healthcare access, and vaccination coverage [7,8].

Primary testing for HR-HPV is a globally recommended cervical cancer screening strategy [9]. The incorporation process is ongoing, and countries are currently at different stages of adoption [10–12]. Several HPV tests that allow genotyping at distinct levels, from limited (partial) to extended, have been approved by the Food and Drug Administration (FDA). Two commercially available assays that provide extended genotyping have been approved for primary HPV screening in women to assess the risk of cervical precancer and cancer, with the most recent approval in 2023 [13]. At the beginning of the 2025 year, the guidelines for applying the results of extended genotyping were revealed by the American Society for Colposcopy and Cervical Pathology. Recommendations apply only for the assay with approval received in 2018, for other assays guidelines may be developed as data allowing risk calculation become available [14].

Despite advancements in screening and vaccination programs in Poland, cervical cancer remains a significant public health concern, contributing to substantial morbidity and mortality rates among women [15,16]. Therefore, understanding the prevalence and genotype distribution of HR-HPV in the Polish population is essential for developing effective preventive strategies, enhancing screening protocols, and optimizing vaccination programs. The p16/Ki67 dual-stain testing (DS) is an immunocytochemical staining method designed to improve the diagnostic accuracy of detecting HR-HPV-associated cervical precancers. By providing highly sensitive and highly specific biomarkers, this method significantly enhances the efficiency of secondary prevention strategies, including primary HPV-based settings [17–19]. The largest global studies on triage options for HR-HPV results in HPV-based cervical cancer screening, including a Polish investigation on DS [20], have recently been presented by Thrall et al. on behalf of the Clinical Practice Committee of the American Society of Cytopathology [21].

Based on FDA approval, indications for the use of DS include positivity for 12 other HR-HPV test results in primary HPV screening, positivity for 12 other HR HPV test results combined with negative cytology in cotesting, positivity for HPV 16/18 in primary HPV or co-testing, performed in conjunction with the physician’s assessment of patient screening history, risk factors, and professional guidelines [22]. Recently, the Enduring Consensus Cervical Cancer Screening and Management Guidelines also recommended DS to triage HPV-positive results in individuals who underwent primary cotesting and ASC-US or LSIL in cytology was obtained, as well as for triage HPV-positive tests results without genotyping in primary HPV setting [23].

Using a retrospective design, this study evaluates the prevalence and genotype distribution of HR-HPV in a large representative sample of women in Poland. These findings are expected to provide valuable insights into the national burden of HPV infection, contributing to the existing body of knowledge on cervical cancer epidemiology and prevention [24]. By presenting the prevalence rates and genotype distribution of HR-HPV in p16/Ki67 DS results using limited and two types of extended genotyping, this study aims to support evidence-based decision-making and guide the development of medical interventions for public health policies worldwide targeted to an incorporation of the new technologies and methods into cervical cancer screening programs. No previous data exist regarding the prevalence and distribution of HR-HPV across all categories of cytological results combined with p16/Ki67 DS status using limited or extended HPV genotyping, including two distinct types of the latter. Therefore, we conducted the first comprehensive investigation including these new technologies and compared our findings with those of other studies.

## Results

### Overall HR-HPV prevalence and trends observed

In the final study group consisting of 10,218 women, 15.0% (1528/10,218) were HR-HPV-positive (HR-HPV+) cases. Among HR-HPV-positive cases, 4.2% (427/10,218) were HPV 16+, 0.9% (96/10,218) were HPV 18+ and 11.6% (1189/10,218) were non-16/non-18 HR-

HPV+ (HPV HR12+). The HR-HPV prevalence in the following years is shown in Figure 1. During the first four years of our study, the overall HR-HPV prevalence, comprising all types of molecular testing performed, showed a gradual decrease. However, a reverse trend was observed, with a slow increase from that point until the last day of investigation period. The observed decrease might be associated with the larger sample size recorded in years 2018-2019 (Figure 2).

**Figure 1.**
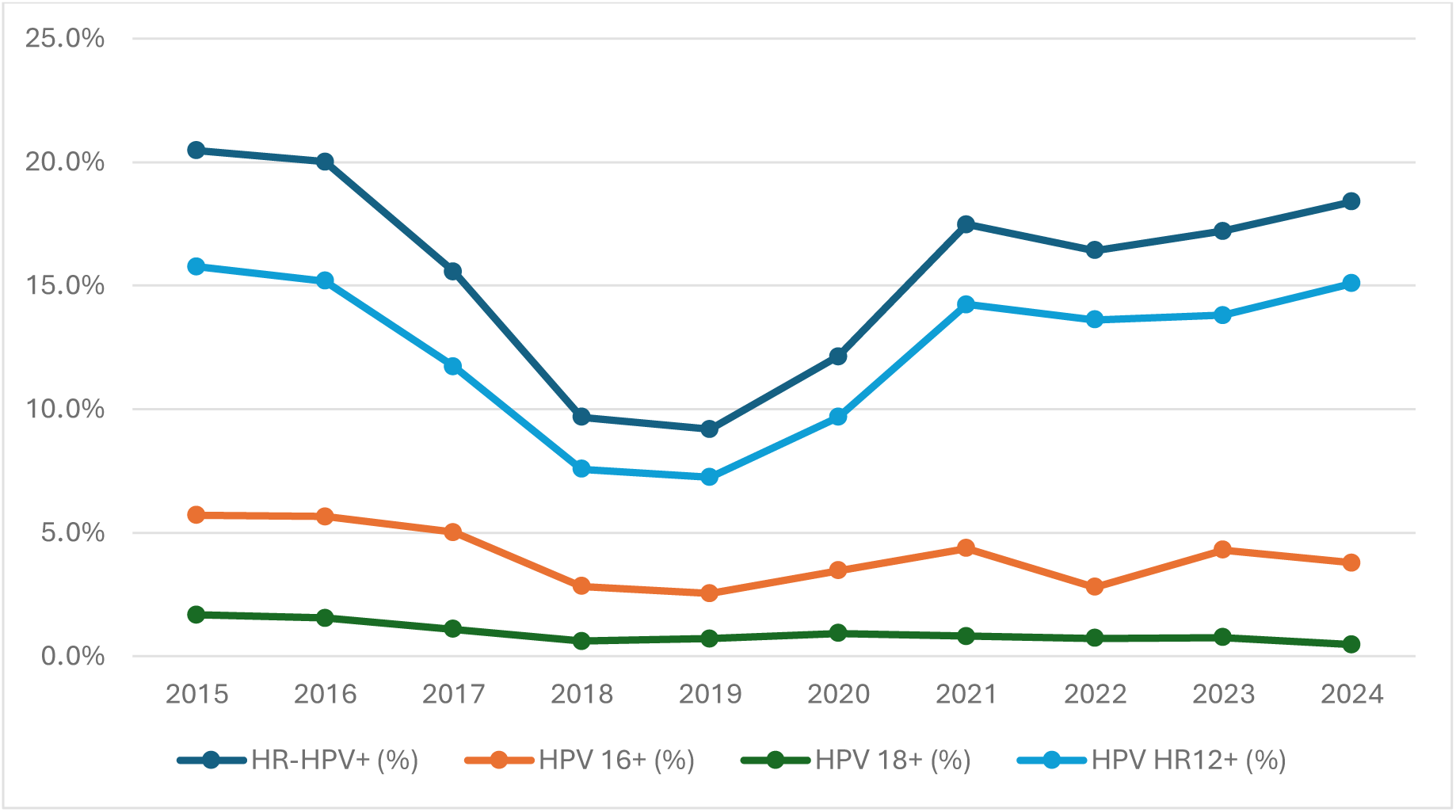
The prevalence of HR-HPV infection. Abbreviations: HR-HPV, high-risk types of human papillomavirus; HR-HPV+, 14 high-risk types of human papillomavirus positive results; HPV 16+, human papillomavirus type 16 positive results; HPV 18+, human papillomavirus type 18 positive results; HPV HR12+, human papillomavirus 12 high-risk types other than types 16 and 18 positive results.

**Figure 2.**
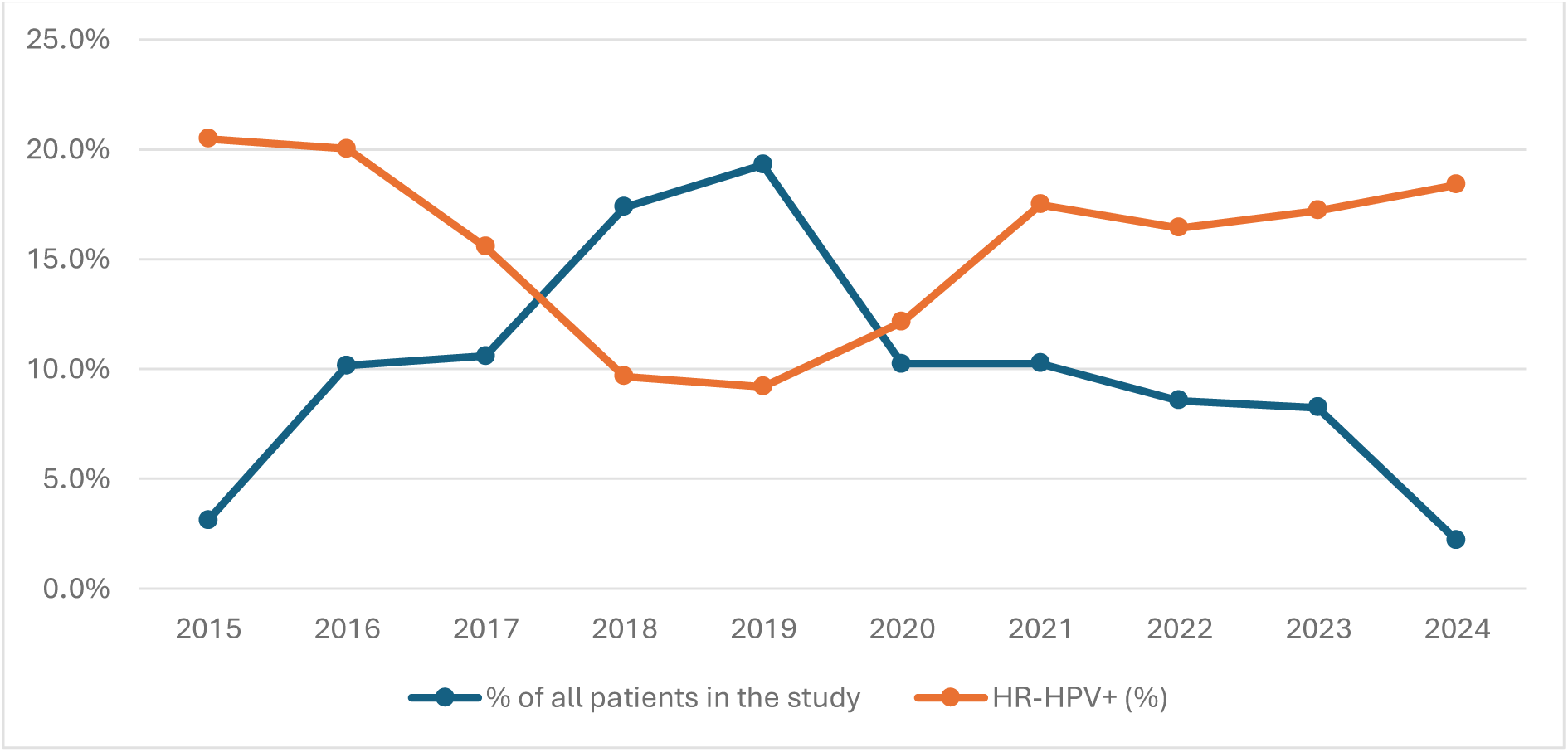
The correlation of number of HR-HPV tests performed per year and HR-HPV prevalence. HR-HPV, high-risk types of human papillomavirus; HR-HPV+, 14 high-risk types of human papillomavirus positive results.

### Limited vs. extended genotyping

HR-HPV testing with limited genotyping was positive in 1,009 cases, with the most common types being HPV HR12 (10.6%) and HR-HPV 16 (4.3%). Molecular tests with extendedgenotyping were HR-HPV-positive in 519 cases, 243 cases the Onclarity assay, and 276 cases using the Alinity assay. This group allowed for extended HR-HPV genotype distribution analysis (Figure 3). The most prevalent genotypes in the Onclarity group were HR-HPV 59/56/66 (5.6%), 16 (4.0%), and 31 (2.8%). The least common were HR-HPV 33/58 (1.0%) and 18 (0.9%). In women tested with Alinity, the set of HR-HPV genotypes with 35/39/51/56/59/66/68 (8.0%) was the most frequently detected, whereas HPV 18 was the least detected (0.4%). The combined prevalence of grouped HR-HPV 33/58 (1.0%) and individual types 31 (2.8%) and 52 (2.1%) in Onclarity group was 5.9%, whereas the corresponding set of genotypes 31/33/52/58 in Alinity group had an identical prevalence of 5.9%. Similarly, sets of genotypes HR-HPV 35/39/68 (2.3%), 59/56/66 (5.6%), and individual HR-HPV 51 (2.2%) in the Onclarity group were detected in 10.1% of women, whereas in the Alinity group the set of genotypes with HR-HPV 35/39/51/56/59/66/68 was detected in 8.0%.

**Figure 3.**
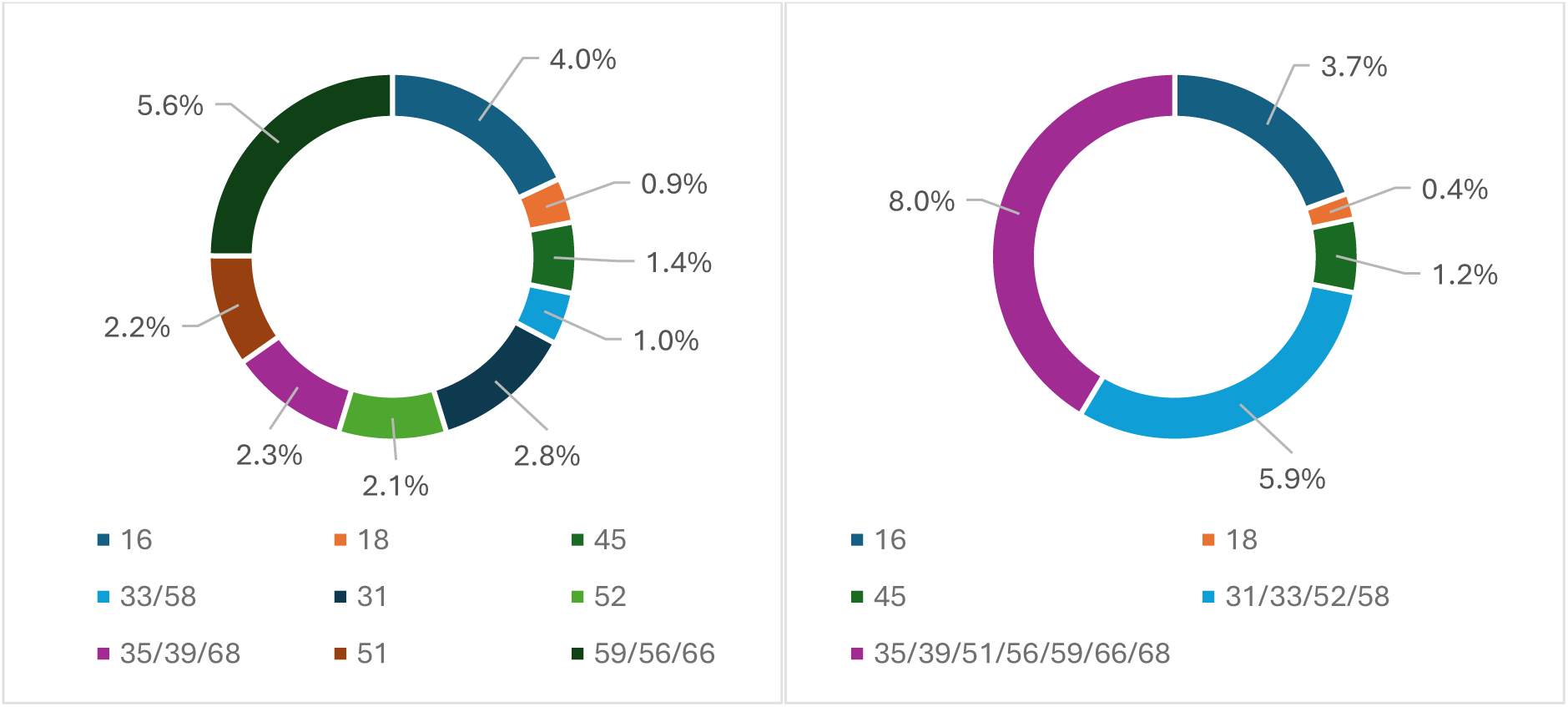
Individual and grouped HR-HPV distribution in Onclarity (on the left) and Alinity (on the right) assays.

The overall HR-HPV prevalence for the results of the Abbott assay was 13.9%, in the Onclarity study group 17.8%, and in the Alinity group 17.2%. There was a statistically significant difference (p<0.0001) in the proportions of positive and negative cases among the three test groups. The variation in the prevalence of HR-HPV between the Abbott group (7250 participants) and the two groups with extended genotyping (2968 participants) may be associated with the difference in sample sizes. The sample size of the first group was nearly 2.5 times larger than the second group. This is also consistent with the lower HR-HPV prevalence observed during the 2018-2019 period, when the study group was at its largest. The differences between the Abbott and Onclarity study group (p=0.00022), and between the Abbott and Alinity groups (p=0.00083) were statistically significant, with a p-value well below the adjusted significance level (0.0167). However, there was no statistically significant difference between the Onclarity and Alinity study groups, as the p-value (0.706) was higher than the adjusted significance level.

### Age-specific HR-HPV prevalence and distribution

The women were divided into selected age groups: <20, 20-24, 25-29, 30-34, 35-39, 40-44, 45-59, 50-54, 55-59, 60-64, and >64 years old. The largest population in the study was in the age range of 35-39 years (1,912), followed closely by the age groups of 30-34 (1,663) and 40-44 (1,643). The smallest sample size was noted in the age group <20 (49). Differences in HR-HPV prevalence and distribution were observed between the groups. Women aged <20, 20-24, and 25-29 had the highest HR-HPV prevalence (34.7%, 26.4%,26.1%, respectively). A decreasing trend in prevalence was noted with increasing age, except for the oldest patients, where a slight increase was observed. In the genotype distribution, the peaks did not fully correspond to prevalence trends. HR-HPV 16 was most prevalent in patients aged 25-29 (8.3%) and 20-24 (8.1%), but was least common in the >64 (1.6%) and 55-59 (1.2%) age groups. HPV HR12 was the most common in the <20 (32.7%) age group and least common in the 60-64 (3.6%) age group. Detailed results are presented in Figure 4.

**Figure 4.**
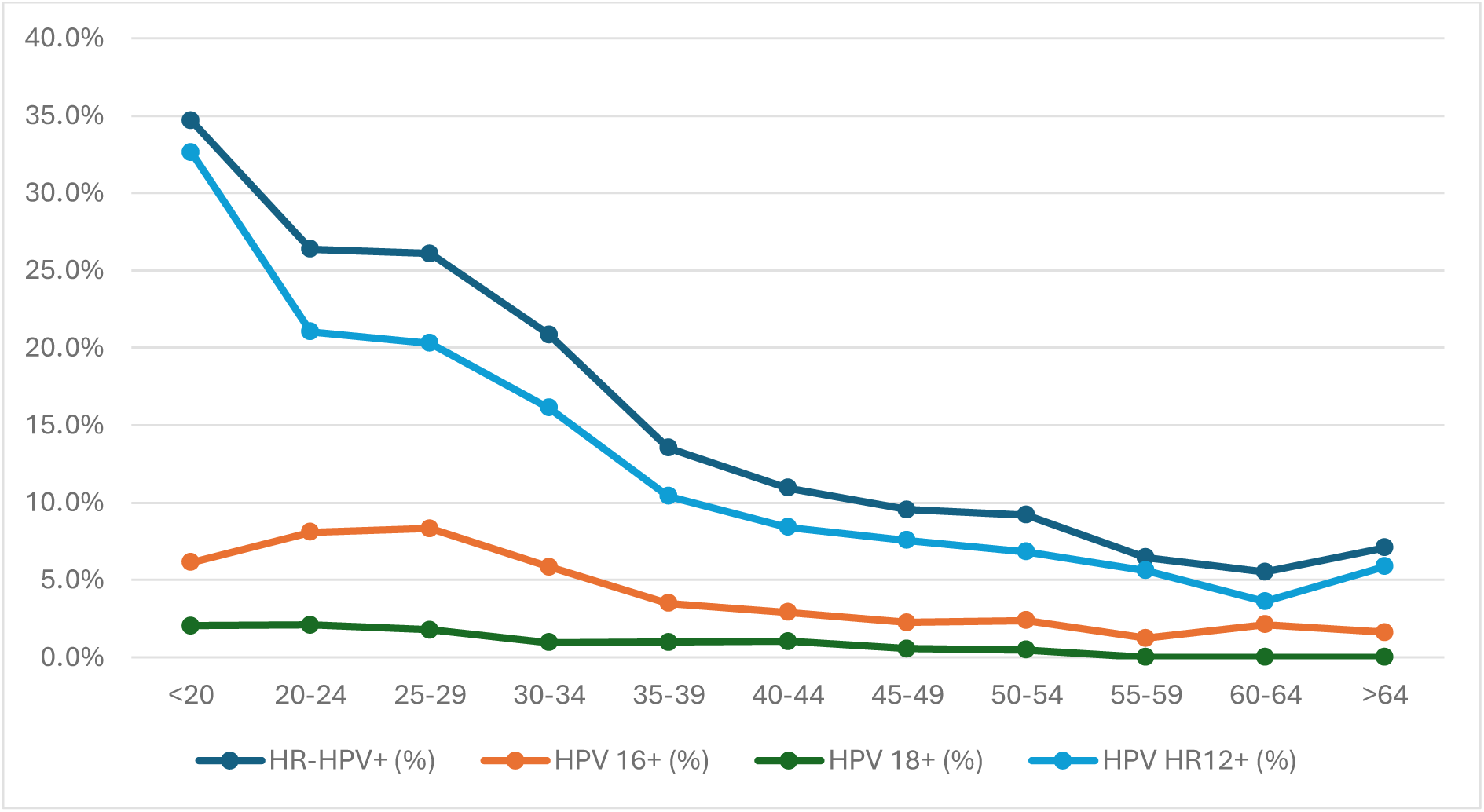
The age-specific prevalence of HR-HPV infection. Abbreviations: HR-HPV, high-risk types of human papillomavirus; HR-HPV+, 14 high-risk types of human papillomavirus positive results; HPV 16+, human papillomavirus type 16 positive results; HPV 18+, human papillomavirus type 18 positive results; HPV HR12+, human papillomavirus 12 high-risk types other than types 16 and 18 positive results.

### Cytology-specific HR-HPV prevalence and distribution

In the final study group, 9,864 women had available LBC results, of whom 12.2% were ASC-US+. The highest HR-HPV prevalence was observed in patients with HSIL (96.7%), ASC-H (85.1%), and LSIL (80.0%). The lowest prevalence was noted in the NILM cases (7.6%). In patients with NILM, ASC-US, LSIL and ASC-H the most frequently detected HR-HPV were HPV HR12, whereas in women with HSIL HR-HPV 16 was the most common. More data are shown in Table 1 (data for 45 cases with AGC, adenocarcinoma and unsatisfactory results are not included in this table).

**Table 1.**
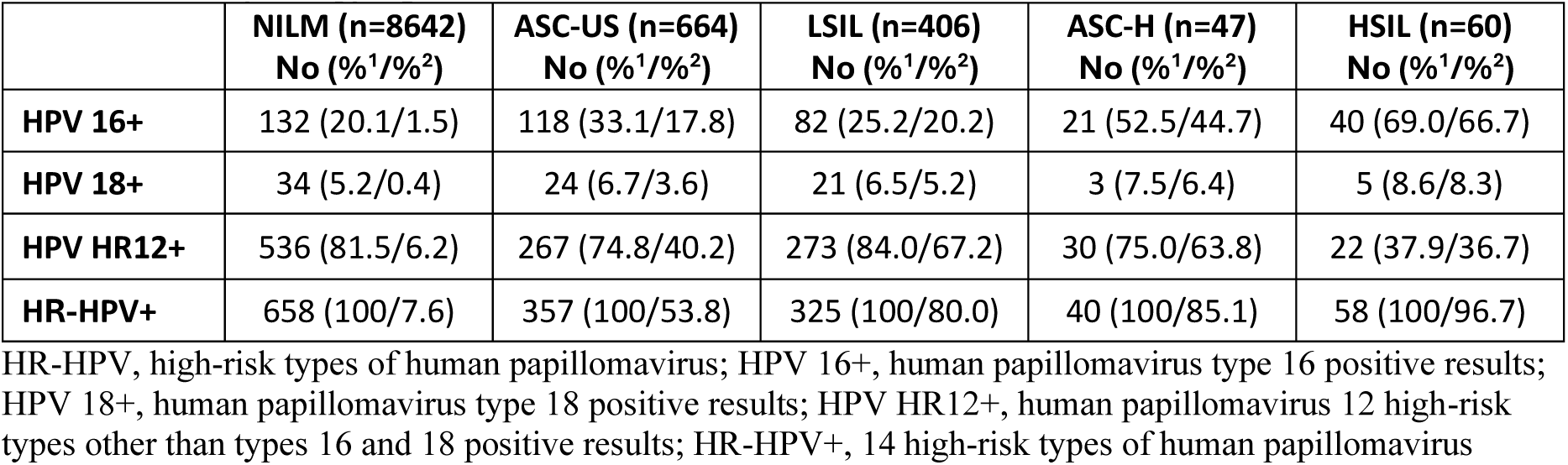
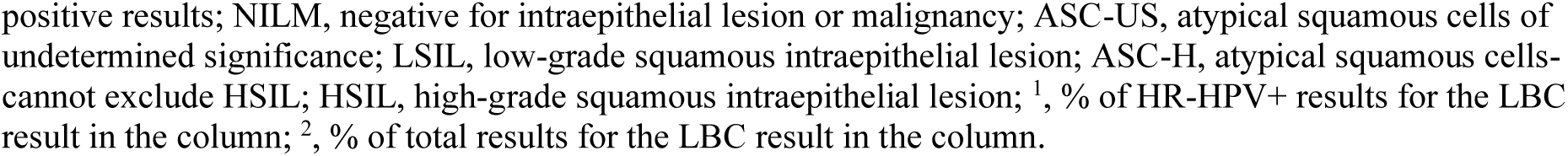
The cytology-specific HR-HPV distribution.

|  | NILM (n=8642)<br>No (%<sup>1</sup>/%<sup>2</sup>) | ASC-US (n=664)<br>No (%<sup>1</sup>/%<sup>2</sup>) | LSIL (n=406)<br>No (%<sup>1</sup>/%<sup>2</sup>) | ASC-H (n=47)<br>No (%<sup>1</sup>/%<sup>2</sup>) | HSIL (n=60)<br>No (%<sup>1</sup>/%<sup>2</sup>) |
| --- | --- | --- | --- | --- | --- |
| HPV 16+ | 132 (20.1/1.5) | 118 (33.1/17.8) | 82 (25.2/20.2) | 21 (52.5/44.7) | 40 (69.0/66.7) |
| HPV 18+ | 34 (5.2/0.4) | 24 (6.7/3.6) | 21 (6.5/5.2) | 3 (7.5/6.4) | 5 (8.6/8.3) |
| HPV HR12+ | 536 (81.5/6.2) | 267 (74.8/40.2) | 273 (84.0/67.2) | 30 (75.0/63.8) | 22 (37.9/36.7) |
| HR-HPV+ | 658 (100/7.6) | 357 (100/53.8) | 325 (100/80.0) | 40 (100/85.1) | 58 (100/96.7) |

HR-HPV, high-risk types of human papillomavirus; HPV 16+, human papillomavirus type 16 positive results; HPV 18+, human papillomavirus type 18 positive results; HPV HR12+, human papillomavirus 12 high-risk types other than types 16 and 18 positive results; HR-HPV+, 14 high-risk types of human papillomavirus
positive results; NILM, negative for intraepithelial lesion or malignancy; ASC-US, atypical squamous cells of undetermined significance; LSIL, low-grade squamous intraepithelial lesion; ASC-H, atypical squamous cells-cannot exclude HSIL; HSIL, high-grade squamous intraepithelial lesion; <sup>1</sup>, % of HR-HPV+ results for the LBC result in the column; <sup>2</sup>, % of total results for the LBC result in the column.

In HPV16+ cases, 33.2% of patients had NILM cytologic result, 29.6% had ASC-US, 20.6% had LSIL, 10.1% had HSIL, and 5.3% had ASC-H. Among HPV18+ patients, 37.8% had NILM cytologic results, 26.7% had ASC-US, 23.3% LSIL, 5.6% HSIL, and 3.3% had ASC-H. The HPV HR12+ results were 47.3% of, 24.1%, 23.6%, ASC-US, 2.6%, ASC-H, and 1.9% in cytological NILM, LSIL, ASC-US, ASC-H and HSIL, respectively.

In the Onclarity group, HR-HPV 59/56/66 set was the most prevalent in women with NILM, ASC-US and LSIL cytology results. In women with cytological HSIL, HPV 16 was the most common HR-HPV detected. Similar results were observed in the Alinity group, with the HR-HPV 35/39/51/56/59/66/68 set being the most common in NILM, ASC-US and LSIL cytology results, while HPV 16 was predominant in cytologic HSIL cases. Detailed data for patients with HR-HPV+ results are presented in Figure 5 (data for the ASC-H category are not presented because of the relatively small sample size in the Onclarity and Alinity study groups).

**Figure 5.**
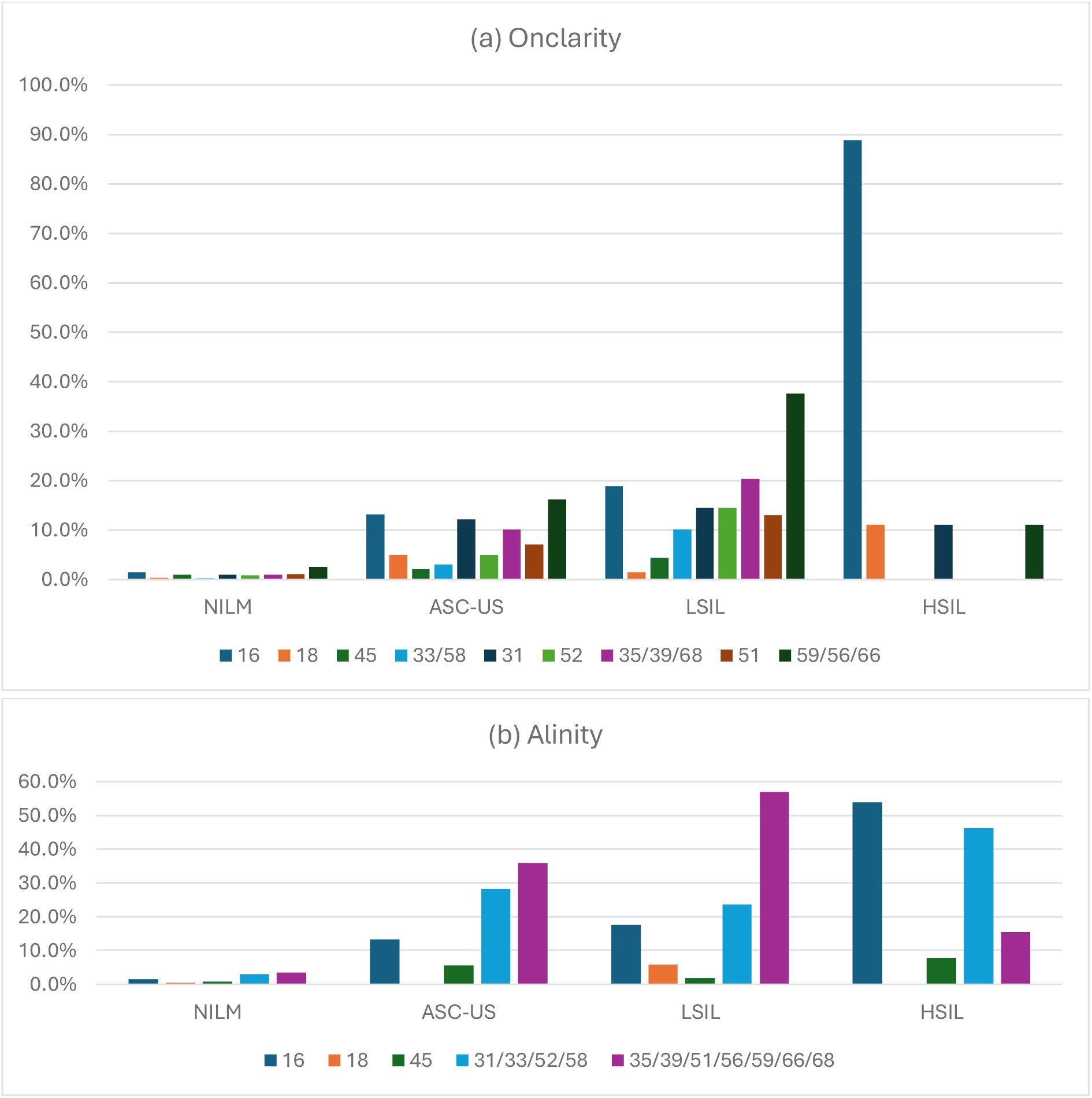
The cytology-specific HR-HPV distribution in women with Onclarity (a) and Alinity (b) assays performed. Abbreviations: HR-HPV, high-risk types of human papillomavirus; NILM, negative for intraepithelial lesion or malignancy; ASC-US, atypical squamous cells of undetermined significance; LSIL, low-grade squamous intraepithelial lesion; HSIL, high-grade squamous intraepithelial lesion.

### DS-specific HR-HPV prevalence and distribution

In the final study group, 1,037 patients underwent DS testing with available results, of whom 844 (81.4%) were HR-HPV-positive. In this group, 35.5% of the women were DS-positive and 64.5% were DS-negative. In 46.8% of HPV16-positive patients, 38.9% of HPV18-positive and 31.9% of HPV HR12-positive cases had positive DS status (53.2%, 61.1% and 68.1% were DS-negative, respectively). The values of ASC-US+ cases for the corresponding HR-HPV-positive subgroups presented in Figure 6 were significantly higher: 66.3% for HPV16+ (p<0.0001), 63.0% for HPV18+ (p=0.0028), and 51.5% for HPV HR12+ (p<0.0001).

**Figure 6.**
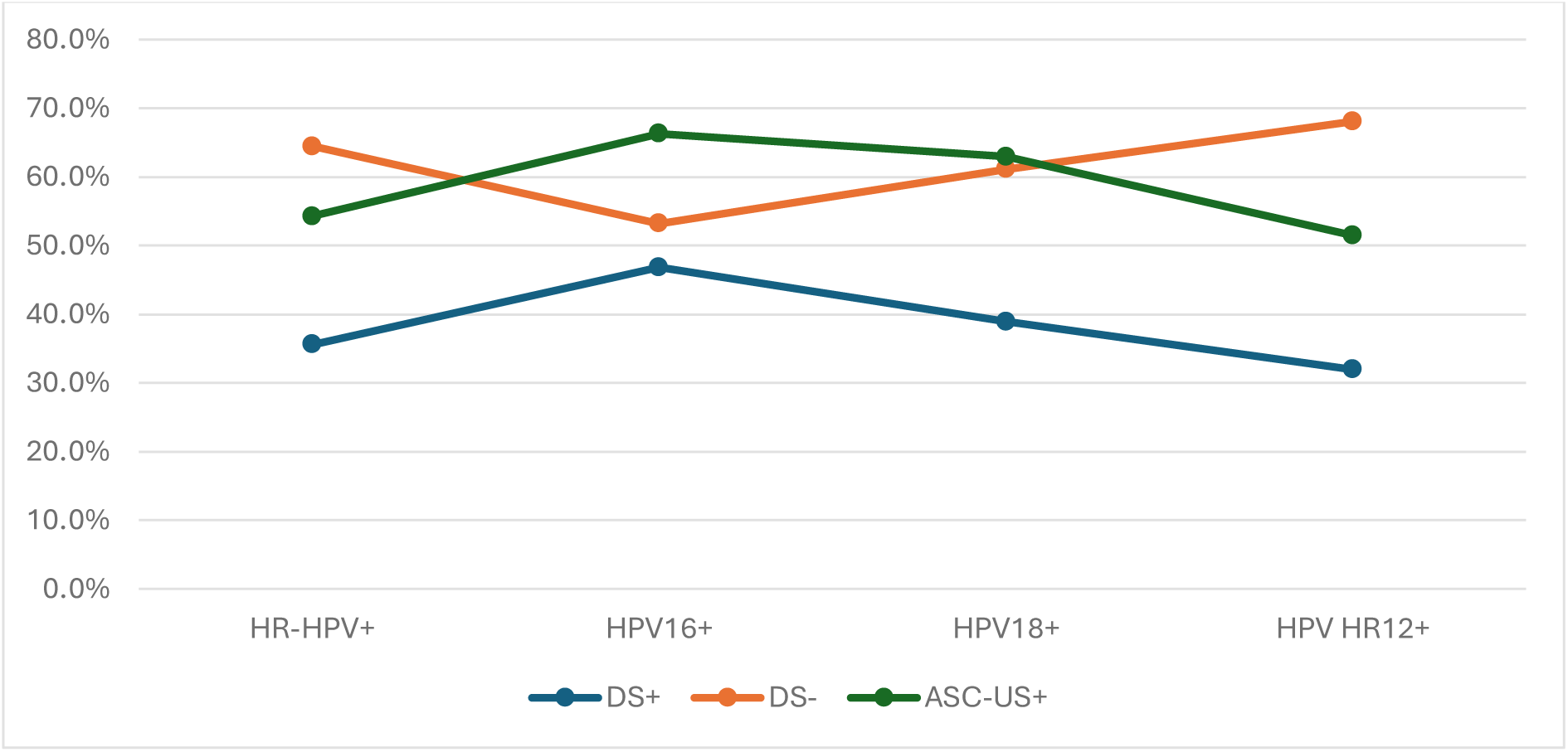
The comparison of DS and LBC results in HR-HPV-positive patients. Abbreviations: HR-HPV, high-risk types of human papillomavirus; HR-HPV+, 14 high-risk types of human papillomavirus positive results; HPV 16+, human papillomavirus type 16 positive results; HPV 18+, human papillomavirus type 18 positive results; HPV HR12+, human papillomavirus 12 high-risk types other than types 16 and 18 positive results; DS, p16/67 dual-stain testing; ASC-US+, atypical squamous cells of undetermined significance or worse; +, positive; –, negative.

In the Onclarity group, 114 women were HR-HPV-positive and had DS performed, while in the Alinity group, 52 women met the same criteria. In the Onclarity group, the highest DS positivity was observed in HR-HPV 33/58 (100.0%) and 31 (58.8%), and the lowest in HR-HPV 45 (18.2%), 18 (25.0%) and 59/56/66 (28.9%). In the Alinity group, the highest DS positivity was noted in HR-HPV 16 (66.7%) and set of genotypes HR-HPV 31/33/52/58 (58.8%), with the lowest observed in the same genotypes as in the Onclarity group. The detailed data for the extended genotyping results are presented in Figure 7.

**Figure 7.**
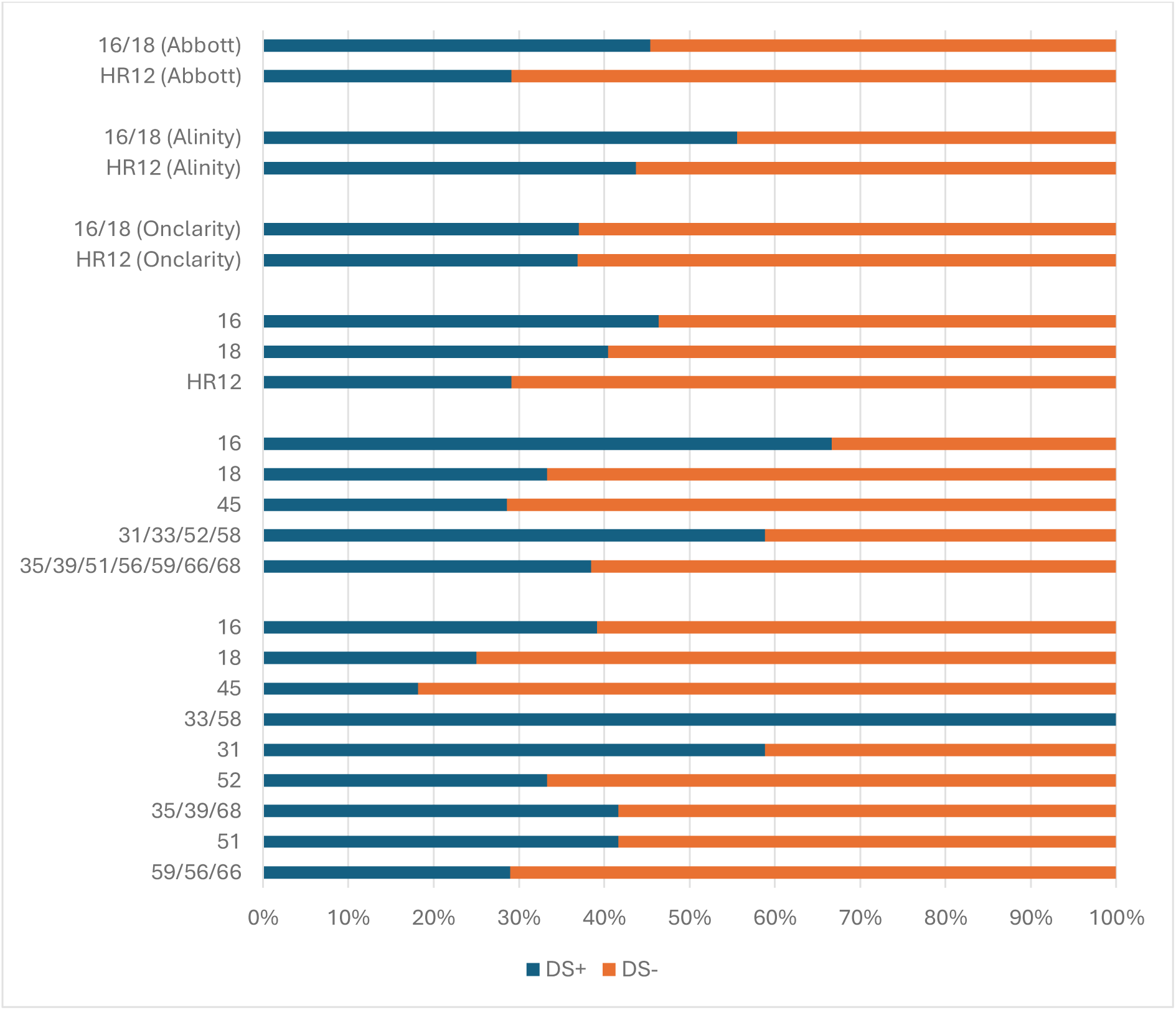
The DS results in HR-HPV-positive patients using Abbott assay (first from the top and third from the bottom), Alinity (second from the top and second from the bottom) and Onclarity assay (third from the top and first from the bottom). Abbreviations: HR-HPV, high-risk types of human papillomavirus; DS, p16/67 dual-stain testing; +, positive; –, negative; 16/18, human papillomavirus types 16 and/or 18; HR12, human papillomavirus 12 high-risk types other than types 16 and 18.

## Discussion

This is the first study to determine the prevalence and genotype distribution of HR-HPV in all categories of cytological results combined with p16/Ki67 DS status in limited genotyping for HPV 16 and/or 18 (the Abbott study group) and for two types of extended genotyping characterizing different individual HPV genotypes or sets of genotypes detected (the Alinity study group; the Onclarity study group). Our findings indicate a significant prevalence of HR-HPV in the investigated population, with notable variation in the genotype distribution. The overall prevalence of the HR-HPV genotypes in our study was 15.0%. HR-HPV 16 was detected in 4.2% of the patients, HR-HPV 18 in 0.9%, and HPV HR12 in 11.6%. The most common HR-HPV types in the Abbott group were other HPV HR12 (10.6%) and HPV 16 (4.3%). In the Onclarity group, HR-HPV 59/56/66 (5.6%), HR-HPV 16 (4.0%) and HR-HPV 31 (2.8%) were the most prevalent genotypes or sets of genotypes. In the Alinity group, the most detected individual or set of HR-HPV genotypes was 35/39/51/56/59/66/68 (8.0%), 31/33/52/58 (5.9%) and 16 (3.7%), with the highest prevalence observed in younger patients (26.1-34.7%). In addition, HR-HPV 16 was the most common HPV type in the younger groups (8.1-8.3%). In women with LBC, HR-HPV was the most prevalent in HSIL (96.7%), which was unsurprising. In the NILM cases, only 7.6% of the patients were HR-HPV-positive. In the NILM, ASC-US, LSIL, and ASC-H cytology results, HPV HR12 was the most frequently detected type (in the Onclarity group, the set of HR-HPV 59/56/66 was the most common in these cytology cases), and in HSIL cases, HR-HPV 16 was predominant. In the HR-HPV-positive group, 35.5% of the women were DS-positive (46.8% of HPV16-positive patients, 38.9% of HPV18-positive, and 31.9% of HPV HR12-positive cases) and 64.5% were DS-negative. The values for ASC-US+ cases were significantly higher: 66.3% (positive for HPV 16), 63.0% (positive for HPV 18), and 51.5% (positive for other HPV HR12). The only clinically important significance for practical management is the difference between patients positive for other HPV HR12 test results with a positive DS status (31.9% for HPV HR12+ DS+) and patients positive for other HPV HR12 test results with abnormal cytology (51.5% for HR12+ ASC-US+). According to most available national guidelines, which recommend molecular tests with limited genotyping for HPV 16 and 18, cases positive for HPV 16 and/or 18 do not require triage and are directly referred for colposcopy [27–30].

The overall HR-HPV prevalence in our study was consistent with findings reported in other European countries [31,32]. Galati et al. reported a comparable prevalence rate was reported, emphasizing the widespread nature of HR-HPV infections across different regions of Europe (14.4% vs. 15.0% in our study) [31]. However, our investigation revealed a slightly lower prevalence compared to Northern European countries such as Sweden and Denmark have been noted, which might be associated with differences in sexual behavior, screening practices, and population demographics (15.0% vs. 18.1%) [32]. The age-specific HR-HPV prevalence was most similar to that observed in other European studies (Nordic countries and Spain), with lower values than in Nordic countries in patients below 30 years old and higher values than in Spain in women below 40 years old [32,33]. In older age groups, the results were nearly equivalent. The results of our study align with other Polish data [34], although they were slightly lower in both overall and HPV 16 prevalence. The most notable differences were observed in the age range 35-44 years. In comparison to a Chinese study, the HR-HPV prevalence was much lower in most of our study groups [35], as well as to another Polish data, reported by Glinska et al. [36] (39% vs. 15.0% in our investigation), likely due to a selected high-risk cohort. The observed overall prevalence was slightly higher than that in the USA data (15.0% vs. 9.8%/10.5%) [37,38]. The age-specific HR-HPV prevalence in our study was close to the similar five-year age groups in the study by Trama et al., except for three groups, where the differences were approximately 10% (26-30/31-35/36-40 – 15.2%/10.8%/6.4% in Trama et al. [37]; 25-29/30-34/35-39 – 26.1%/20.9%/13.5% in our study). The prevalence in other age groups was only slightly higher in our study, most likely due to the younger age groups. The age-specific HR-HPV prevalence was also a slightly higher than that reported by Wright et al., with the largest differences observed in the 30-39 age groups, but less than 10% [38].

Regarding HR-HPV genotype distribution (from the most detailed genotyping performed in our study, the Onclarity group with extended genotyping), HPV 16 was the most prevalent single genotype in our cohort, aligning with global trends that identify this genotype as the most common and oncogenic (4.0% vs. 5.9%, 4.9%, 5.6%, 3.4%, and 3.5%) [31, 32, 39, 40]. Our data showed that the set of genotypes HR-HPV 59/56/66 accounted for a significant proportion of the infections. Additionally, our results demonstrated a similar prevalence of other HR-HPV types, including 31 (2.8% vs. 2.8%/2.9%) [31,32], 51 (2.2% vs. 2.1%) [31], 52 (2.1% vs. 2.8%) [32], and 45 (1.4% vs. 0.8%) [31]. These trends are consistent with the findings of other European studies. In comparison to a study from the USA [37], the prevalence rates for HR-HPV 16 (1.2% vs. 4.0%), 31 (0.5% vs. 2.8%), and the set of HR-HPV 59/56/66 (2.0% vs. 5.6%) genotypes were higher than those in our study. However, HR-HPV 18, 45, 51, 52, 33/58 and 35/39/68 showed similar results. Individually or in sets, HR-HPV 52, 33/58 and 35/39/68 were more prevalent in the recent Chinese study than in our results, but genotypes HR-HPV 45 and 31 were less common [41]. The prevalence rates of HR-HPV 16, 18, 51, and 59/56/66 were similar. These discrepancies could suggest regional variations in genotype distribution, possibly influenced by differences in the local epidemiology of HPV, host genetic factors, or environmental factors. We were not able to compare the prevalence of individual genotypes 33, 35, 39, 56, 58, 59, 66, and 68, as the Onclarity assay provided only grouped results.

The cytology-specific HR-HPV prevalence was similar to that reported by Wang et al. [35]. Comparing the same results to a study from the US, our findings were relatively higher for all parameters, with the most similar results in NILM cytology [37]. Furthermore, our cytology-specific results were higher for HPV HR12+ women with NILM, ASC-US, LSIL and ASC-H cases than in the study by Wentzensen et al. [42]. The results for the cytological HSIL group were very similar (36.0% vs. 37.9% in our study). The overall HR-HPV prevalence in ASC-H cases was slightly higher in our study than in the recent study by Sun et al. (85.1% vs. 78.5%), with type 16 (44.7% vs. 33.8%) and 18 (6.4% vs. 4.7%) more common and HPV HR12 (excluding the cases with types 16 and 18 co-infection; 36.2% vs. 39.1%) less prevalent [43].

The percentage of DS-positive cases among HPV HR12-positive women in our study was lower than reported by Wright et al. (31.9% vs. 47.5%) [44]. The 100% DS positivity for grouped HR-HPV 33/58 in the Onclarity group may be associated with the relatively small number of cases with HR-HPV genotypes that had performed the DS test in the Onclarity group. Our study highlights the importance of ongoing surveillance and monitoring of the HR-HPV genotype distribution. The introduction of HPV vaccination programs is expected to alter the prevalence of vaccine-covered genotypes, potentially leading to an increase in non-vaccine high-risk genotypes. Continuous evaluation of genotype prevalence is crucial in assessing the long-term impact of vaccination and in guiding future vaccine development strategies. This study has several strengths. One of the most significant findings is the analysis of one of the largest numbers of HR-HPV, LBC, and DS results retrieved from real-world clinical practice, which is crucial for informing screening guidelines. Additionally, it includes a wide age range of participants and provides insights into the results of screening tests for opportunistic cervical cancer. Another strength is the joint analysis of HR-HPV, LBC, and DS, with all LBC and DS evaluated by a qualified gynecological cytopathologist providing a standardized gynecological cytopathology evaluation for all investigated screening test results. Colposcopic specimens were standardized for all women in each being reviewed by a gynecologic pathologist using p16 immunohistochemistry to increase the effectiveness of interpretations. Colposcopy, cervical biopsy procedures and endocervical sampling were performed by qualified gynecologists certified and audited by a professional society. However, the study has potential limitations, such as being conducted in a single private fund-based center, operating in a relatively small area, and covering a homogeneous study population. Further multicenter studies are required, to confirm our results.

## Conclusions

In conclusion, the prevalence and genotype distribution of HR-HPV types among women in Poland aligns with broader European and USA patterns, with some regional distinctions.

These findings underscore the critical need for continued HR-HPV surveillance, public health education, and vaccination efforts to mitigate the burden of HPV-related diseases in the era of HPV-based screening and the related entry of new molecular limited or extended genotyping or biomarker DS testing. Future research should focus on longitudinal studies to monitor the impact of vaccination programs and to explore the reasons for regional variations in HR-HPV genotype distribution. A more combined approach, implementing new technologies, enables healthcare providers to make more informed decisions about patient care, ultimately aiding in the early detection and prevention of cervical precancer and cancer, leading to a novel capability to enhance national screening programs.

## Materials and Methods

### Study population

In this study, 32.724 cervical cancer liquid-based screening (LBS) test results were retrospectively investigated. The samples were obtained from opportunistic private founds-based cervical cancer screening conducted at the Corfamed Woman’s Health Center, one of a major private outpatient gynecological clinic located in an urban area in Poland. The analysis was based on virologic, cytologic and immunocytochemical test results retrieved from the Center’s electronical database from August 2015 to March 2024. This study included the following initial screening test results: 15.856 for HR-HPV, 15.195 for LBC, and 1.673 for DS. During the study period, two different screening models were implemented: primary cytology with reflex HR-HPV, and primary cotesting. The reflex HR-HPV test was performed in cases with atypical squamous cells of undetermined significance (ASC-US) or low-grade squamous intraepithelial lesions (LSIL) detected on primary cytology screening. DS was applied for all HR-HPV-positive results, for ASC-US or LSIL in cytology, and in cotesting-based screening models, in accordance with Polish national guidelines [25]. The LBS tests were performed on a diverse group of women aged 16-92 years, with an average age of 41.02 years. The final study group consisted of 10.218 patients. The socioeconomic status of the participants predominantly fell within the middle-to-upper socioeconomic strata. The educational background of the participants was notably high, with all having completed at least secondary education and the majority holding higher education degrees. Eligibility criteria were designed to enroll non-pregnant participants representative of a real-life screening population who had all the investigated screening tests available, including HR-HPV status, cytology, and DS, and who underwent standardized medical procedures. These procedures included management with abnormal screening test results and follow-up monitoring to determine the controlled disease ascertainment. The primary exclusion criteria included hysterectomy, current pregnancy, history of treatment for cervical intraepithelial lesions or cancer, current cancer, or missing data. Secondary exclusion criteria were as follows: cytologic or DS reports interpreted by an unqualified or non-gynecological cytopathologist, and the use of HR-HPV molecular assays different from those specified in the enrolment criteria. Primary and secondary exclusion criteria were consistently applied across all enrollment protocols. The data used in this study were derived from routine cervical cancer screening. Therefore, selected or full clinical data were available and unblinded to individual physicians or specialists performing the screening and were involved in the investigation. For instance, gynecologists performing screening tests, cytopathologists evaluating cytology and DS slides, and laboratories conducting molecular HR-HPV testing all had access to relevant to their level of clinical information, such as patient identification data and clinical context. Other researchers involved in the study had access to unblinded clinical information including the number of study participants and subgroups, patient age, screening test results for each group, and type of molecular assay used. Randomization was not applicable to this study owing to its retrospective design, as it was not a clinical trial. The study was conducted in accordance with recognized ethical guidelines (International Ethical Guidelines for Biomedical Research Involving Human Participants, the Declaration of Helsinki, and applicable local regulations). The study was approved by the Ethics Committee (ID: 118.6120.36.2023).

### HR-HPV molecular testing with limited or extended genotyping

HR-HPV detection was performed using one of the three following assays: the Abbott RealTime High Risk HPV molecular in vitro PCR test (Abbott Molecular, Des Plaines, IL, USA), BD Onclarity HPV Assay (Becton Dickinson, Franklin Lakes, NJ, USA), or Alinity m HR HPV Assay (Abbott Molecular, Des Plaines, IL, USA), all in accordance with the manufacturer’s instructions. The first assay detects 12 types of high-risk HPV DNA (31, 33, 35, 39, 45, 51, 52, 56, 58, 59, 66, and 68) in one group, and identifies HPV 16 and 18 independently (limited genotyping). The second assay detects individually HPV 16, 18, 45, 31, 52, and 51 and reports sets of the following HR-HPV genotypes: 33/58, 35/39/68 and 59/56/66. The Alinity HR HPV Assay detects 16, 18, and 45 separately and classifies other HR-HPV types, into two sets of genotypes: 31/33/52/58 and 35/39/51/56/59/66/68. Both the Onclarity and Alinity assays represent extended genotyping beyond HPV 16 and 18, targeting simultaneous detection of more than one viral genotype. All screening samples were collected using a Cervex-Brush device (Rovers Medical Devices, the catalog number: ROV 380100331). All screening tests (HR-HPV, LBC, and DS) were performed in two separate laboratories: Abbott Real Time and Alinity with LBC SurePath and DS in one laboratory and Onclarity assay with LBC and DS in the other.

### Liquid-based cytology

The external laboratories prepared liquid-based SurePath slides using an automatic BD PrepStain Slide Processor (RRID:SCR_026336) and BD Totalys SlidePrep (RRID:SCR_026341) according to the manufacturer’s instructions. All the residual cervical samples were stored in the laboratories for 1-3 months. A gynecological cytopathologist evaluated all cytology samples based on the Bethesda 2014 system, with knowledge of the HR-HPV status of the samples (an informed cytology approach). Quality and control procedures adhered to the standards of US laboratories accredited by the College of American Pathologists [26].

### p16/Ki67 dual-stain testing

A CINtec PLUS Detection Kit, Roche, Cat# 10215348001 (RRID:AB_3675723) was used for dual immunocytochemical staining. This kit contains a cocktail of two primary antibodies targeting human proteins: (1) a monoclonal mouse antibody (clone E6H4) directed against the cell cycle regulator p16^INK4a^ protein and (2) a recombinant rabbit primary antibody (clone 274-11AC3V1) directed against the proliferation marker Ki-67 protein. Immunocytochemical detection of p16 and Ki-67 proteins in cytological specimens was performed using an automated Ventana BenchMark XT system (RRID:SCR_026335), with 3,3’-diaminobenzidine tetrahydrochloride (DAB) and Fast Red as signal detection chromogens, according to manufacturer’s instructions. The procedure was carried out as previously described [18–20, 26] using residual cellular samples from the original SurePath vials after cytology and/or HPV testing. p16/Ki67 testing was not performed if previously assessed cytology was reported to be inadequate. Immunoprofile assessment was conducted by a qualified and experienced gynecological cytopathologist who was blinded to the cytology and HR-HPV test results. p16/Ki67 slides were categorized as positive, negative, or unsatisfactory. A slide was considered positive if at least one epithelial cell exhibited concurrent red nuclear staining for Ki67 and brown cytoplasmic staining for p16. In cell clusters, a positive result was characterized by strong, diffuse p16 staining, and the identification of at least one cell on the periphery with nuclear Ki67 positivity and cytoplasmic p16 expression. Slides were defined as negative when no immunostaining was detected or when staining for either p16 or Ki67 was observed within the epithelial cell.

## Statistical analysis

The screened women were categorized into 11 groups. The distribution of HR-HPV was described using indicators such as prevalence and type-specific HPV positivity rates (%). Prevalence was defined as positive test results for HR-HPV. Statistical analyses were conducted to compare the distribution of positive and negative results across different test groups. The chi-square test (χ^2^) was used to determine the overall associations, followed by pairwise comparisons using the chi-square test with Bonferroni correction. Additionally, binomial test (McNemar’s) was applied to assess the significance of the differences between DS-positive and LBC-positive (ASC-US+) results in HR-HPV-positive cases. *p* < 0.05 was considered statistically significant. This study was a retrospective analysis, and the sample size was determined by the availability of all eligible data to maximize the dataset for analysis. Therefore, formal power calculations are not required. All statistical analyses were conducted using Microsoft Excel 365 (RRID:SCR_016137) and licensed 2015 PQStat Statistical Calculation Software (RRID:SCR_026338).

## Data Availability Statement

The data generated in this study are available within the article. All other raw data are available upon reasonable request from the corresponding author.

## Ethics Statement

The study was approved by the Ethics Committee of Jagiellonian University (opinion ID 118.6120.36.2023). The study was conducted in accordance with the principles of Declaration of Helsinki. Informed consent was not obtained because of the retrospective design of this study.

## Author Contributions

Conceptualization, M.T., and M.M.; Collecting of data, K.M., and M.T.; Data curation, K.M. and M.M.; Analysis of data, M.M., M.T., and K.M.; Interpretation of data, M.T., K.M. and M.M.; Investigation, M.M., M.T.; Writing draft, M.T., K.M. and M.M.; Draft editing, M.T., K.M. and M.M.; Supervision and review, R.J and A.H.; All authors have read and agreed to the published version of the manuscript.

